# Ranking hip and knee joint contact forces during high-impact activities in high-functioning adults after hip or knee arthroplasty

**DOI:** 10.64898/2026.02.23.26346712

**Authors:** Bernard X.W. Liew, Jiayu Hu, Zainab Altai, Ahmed Soliman, Leiming Gao, Stephen McDonnell, Wenxing Guo, Stefan Maas, Nelson Cortes

## Abstract

**Background:** People with hip or knee joint arthroplasties are commonly advised to avoid high-impact physical activities, despite increasing demand to return to sport and vigorous exercise. Current implant testing standards do not reflect real-world loading during high-impact tasks, and few studies have quantified implant loads in high-functioning individuals who have returned to such activities.

**Methods:** High-functioning adults with a total hip arthroplasty (THA, *n* = 11), total knee arthroplasty (TKA, *n* = 4), or unicompartmental knee arthroplasty (UKA, *n* = 3) performed a range of low-to high-impact activities, including walking, running, hopping, countermovement jumps, landings, and change-of-direction tasks. Three-dimensional trunk and lower-limb kinematics and ground reaction forces were collected. Musculoskeletal modelling was used to quantify three-dimensional hip and knee joint contact forces. Linear mixed-effects models were used to rank implant loads across activities and to compare peak resultant joint loads with healthy controls from a prior study.

**Results:** For people with THR, relative to walking, a 45° change of direction generated the highest predicted hip contact force (8.38 BW, 95% CI 7.70-9.06), followed by running and unilateral hopping (all >1.5× walking, *p* < 0.05). Unilateral hopping and running produced the highest predicted knee contact force in TKA and UKA participants (8.0-9.1 BW), and both significantly greater than walking (*p* < 0.05). Compared with healthy controls, THA participants exhibited a lower predicted HCF during walking (–1.58 BW, 95% CI –2.46 to –0.69), but no group differences were observed for running, hopping, or jumping.

**Conclusion:** High-impact activities vary widely in model-estimated hip and knee contact forces. Several tasks were not substantially higher than walking. These data provide a biomechanical basis for evidence-informed activity prescription, regulatory implant testing, and future computational simulation of implant performance under realistic loading conditions.

## Introduction

The demand for hip or knee joint arthroplasties is increasing [1], particularly in the younger populations [2]. Despite significant advancements in implant engineering and surgical techniques [3], people with a hip or knee joint arthroplasty are still advised against participating in vigorous, high-impact physical activities (PA) and sports [4–6]. Current guidance often treats high-impact activity as a single category because task-specific estimates of hip and knee joint loading after arthroplasty are limited.

While observational studies have reported that high-impact PA do not increase joint arthroplasty revision rates [7, 8], clinical study designs cannot easily determine the mechanical load thresholds that balance short– and long-term joint safety and musculoskeletal benefits. Previous studies have shown that PA, like hopping, which places ∼7-8BW of force on the hip and knee, is especially effective for improving femoral neck bone density [9]. This is especially important for people with a hip or knee joint arthroplasty, given that they are at an age where the risk of osteoporosis and sarcopenia is greater [10–12].

Few studies have quantified joint implant loads in patients with a hip or knee joint arthroplasty during high-impact PA. Bergmann and Colleagues reported in vivo hip and knee implant loads during a variety of PA from walking, stairs, to running [13, 14]. They reported a peak resultant hip and knee contact force magnitudes of ∼4.2BW in patients with a total hip and total knee arthroplasty, respectively, whilst running between 1.8-1.9m/s on a treadmill [13, 14]. There was limited reporting of the physical function status of the included participants from the previous studies [13, 14]. Speculatively, these could mean that prior studies have not included patients with a joint arthroplasty who have returned to high-impact activities. This could have precluded investigating implant loads across a wider range of high-impact activities beyond running, like hopping, countermovement jumps (CMJs), and change of direction manoeuvres (CODs) [15].

Being able to return to high-impact PA represents a departure from traditional clinical practice [4–6], which creates a natural inertia for change. Part of this inertia arises because joint implants are not stress-tested against loads that reflect high-impact PA. To address this gap, we estimated and ranked peak model-derived hip and knee resultant joint forces across common high-impact tasks in high-functioning adults after total and partial hip or knee arthroplasties and compared selected tasks with healthy controls.

## Methods

### Participants

Participants with a total hip arthroplasty (THA, n = 11), total knee arthroplasty (TKA, n = 4), or unicompartmental knee arthroplasty (UKA, n = 3), who met the eligibility criteria, were _invited. Adult participants were eligible if they had 1) undergone a THA, TKA, or UKA surgery_ ≥ _1_ year from the point of testing, 2) a self-reported status of a return to high-impact PA (e.g. running, jumping), and 3) not experiencing any pain at the site of the joint arthroplasty. All participants provided written informed consent, and the study received approval from the Health Research Authority Research Ethics Committee (IRAS 327418). Data from healthy control participants came from a previous study [16].

### Experimental protocol

Participants with a joint arthroplasty performed all testing whilst wearing their comfortable running shoes and exercise attire. For the present study, retro-reflective markers were adhered to the trunk and lower extremities of each participant (see Supplementary Figure SM1). Participants performed two testing sessions on the same day (with a two-hour break in between). In the first session (overground trials), marker trajectories were captured using 15 optical motion capture cameras (Vicon Ltd., 200 Hz), while ground reaction forces (GRF) were captured using two in-ground force plates (Kistler, 2000 Hz) positioned side by side. In the second session (treadmill trials), marker trajectories were captured using 9 optical motion capture cameras (Vicon Ltd., 200 Hz), while ground reaction forces (GRF) were captured using an instrumented split (left-right)-belt treadmill (Bertec, 2000 Hz).

Table 1 provides a list of task abbreviations used. For overground walking and running, participants performed at least three good trials at a self-determined speed. For CMJs, participants performed three separate repetitions of maximal and submaximal (50% perceived maximal) effort CMJs, each at the preferred descent depth, with the arms held in a 90° abducted “T” pose. For each jump, each foot was positioned on each force plate, shoulder width apart. For COD, participants performed side-step cutting with their operated limb, at a self-determined approach running speed, across three cut angles (45°, 90°, 180°). For each cut angle, three successful trials were required. For vertical hopping, participants performed the task over four conditions: bilateral or unilateral (operated limb) at their self-determined frequency; and bilateral or unilateral (operated limb) at a fixed moderate intensity of 2.6Hz using an auditory metronome. Participants were required to position their arms in a 90° abducted “T” pose and hop continuously for 10s. A minimum of 1 min rest was provided between tasks to avoid fatigue. For landing, participants performed a bilateral drop-land from steps of height 10cm and 20cm, with their arms abducted to a “T” pose position. They were instructed to land using their self-determined technique. Three successful trials per step height were undertaken.

**Table 1.**
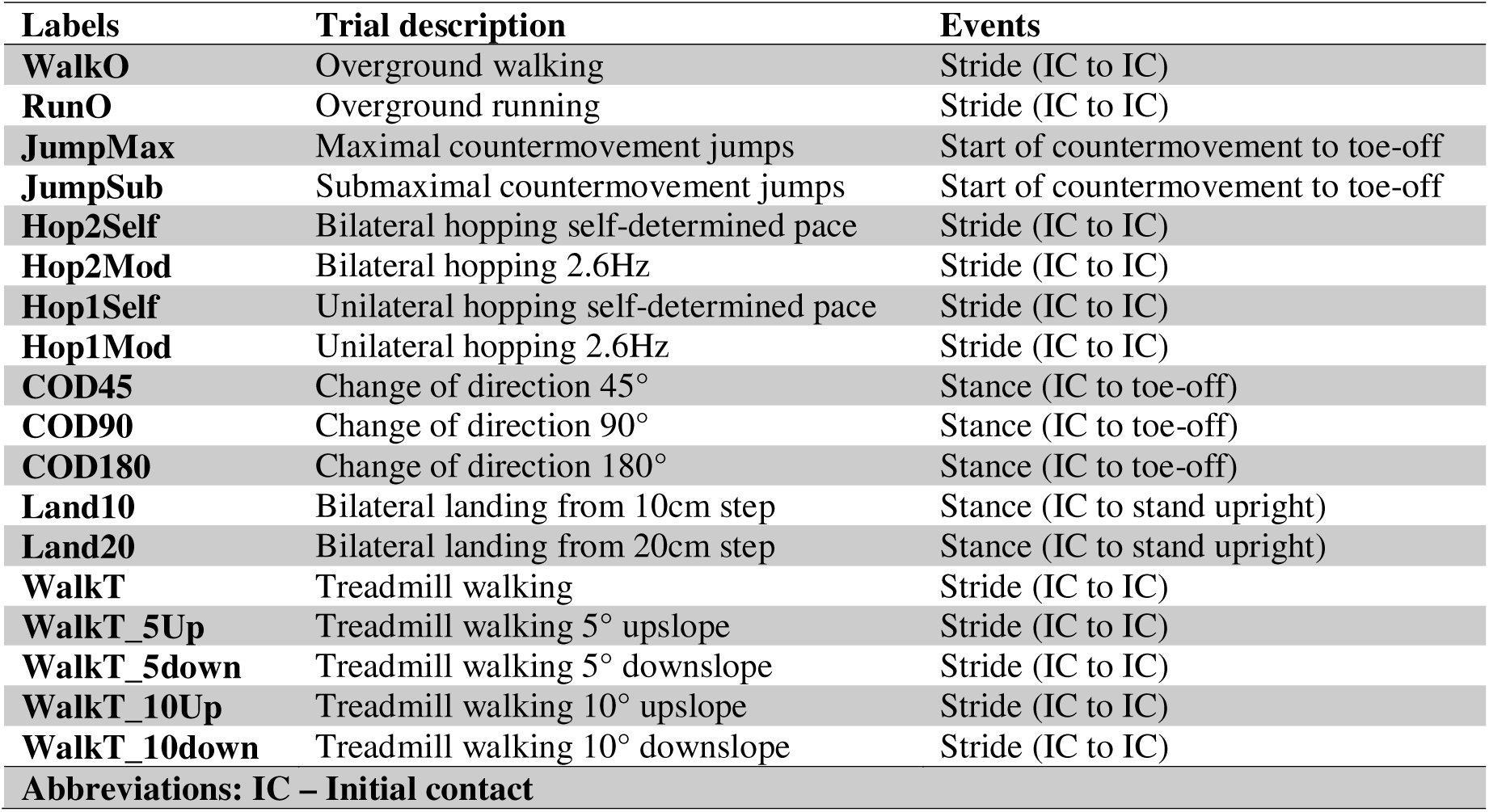
List of abbreviations used to describe the tasks and movement cycle analysed.

For treadmill trials, participants performed walking and running on a level surface at their self-determined speed. To determine the self-determined speed, participants were provided with a practice period. Once the participants had confirmed their speed, the investigator recorded the treadmill speed. Participants then performed walking upslope and downslope at a 5° and 10° incline, at the speed recorded for level-surfaced walking. For all treadmill trials, a body support harness was worn to mitigate the risk of falling, and participants performed 30s of continuous walking or running.

Details of the healthy participant protocol are reported in Altai et al. (2024) [16] and summarised in Table SM1 in the supplementary material. The following movement tasks were common between the two studies and were used for between-group comparisons: overground walking and running, maximal and submaximal CMJs, and bilateral and unilateral hopping at 2.6Hz.

### Musculoskeletal modelling

Marker trajectories were low-pass filtered (2nd-order Butterworth, 5 Hz). GRFs were low-pass filtered (2nd-order Butterworth, 18 Hz). Both were used as input for an inverse dynamics analysis in the AnyBody Modelling System (v8.1, AnyBody Technology A/S, Aalborg, Denmark) [17]. A detailed generic musculoskeletal model, the Twente Lower Extremity Model version 2 (TLEM2), including the trunk [18], based on a cadaveric dataset [19], was scaled to match the height and body segment lengths of each participant based on the marker set collected during a static standing trial [20]. Muscle strength was scaled according to each participant’s body mass. An inverse dynamics analysis based on a third-order polynomial muscle recruitment criterion was then performed to calculate the required muscle forces [21]. The resultant joint contact force at each joint was calculated, normalised to body weight (BW), and the maximal value within the movement cycle was extracted.

To facilitate between-group comparisons, the intensities of the tasks in both studies were quantified and used as a covariate: the average centre of mass (COM) speed (m/s) for walking and running, the average COM approach speed (m/s) for all COD tasks, stance duration (s) for hopping, and the jump height (m) for the CMJ, defined as difference between the peak and the standing height of the COM.

### Statistical analysis

All statistical analyses were conducted using R software (version 4.4.2). We compared the present hip contact force (HCF)’s mean (SD) waveforms to an average 75kg subject walking overground at a self-determined average speed of 1.1 m/s and treadmill running at 1.9 m/s [13]. We also compared the present knee contact force (KCF)’s mean (SD) waveforms to an average 75kg subject walking overground at a self-determined average speed of 1.1 m/s and treadmill running at 1.7 m/s [14].

The dependent variables are the peak resultant HCF and the knee contact force (KCF). For the first aim, the independent variables are the different movement tasks (11 tasks with walking as the reference), different groups (three groups with THA as the reference), a movement-by-group interaction, and a random-subject intercept. For the second study aim, the independent variables are the different groups (four groups with control participants as the reference), participants’ age, tasks’ intensity, and a random-subject intercept. We fitted a linear mixed model, followed by a Type III analysis of variance on the fitted models. For the first aim, post-hoc pairwise task contrasts against walking were computed. For the second aim, pairwise group differences against the control group were computed. We used a False Discovery Rate (FDR) adjusted P-value of <0.05 to determine statistical significance.

## Results

Table 2 provides a summary of the descriptive characteristics of our participants. Table 3 provides a summary of the task intensities performed. The supplementary material contains figures of the average resultant contact force waveforms. For the HCF and KCF during walking, the peak difference between the average modelled forces and Bergmann’s data was 2.36 BW and 1.36 BW, respectively (see Supplementary Material). In running, peak differences with Bergmann’s were 5.02 BW and 4.91 BW for hip and knee contact forces, respectively (see Supplementary Material).

**Table 2.**
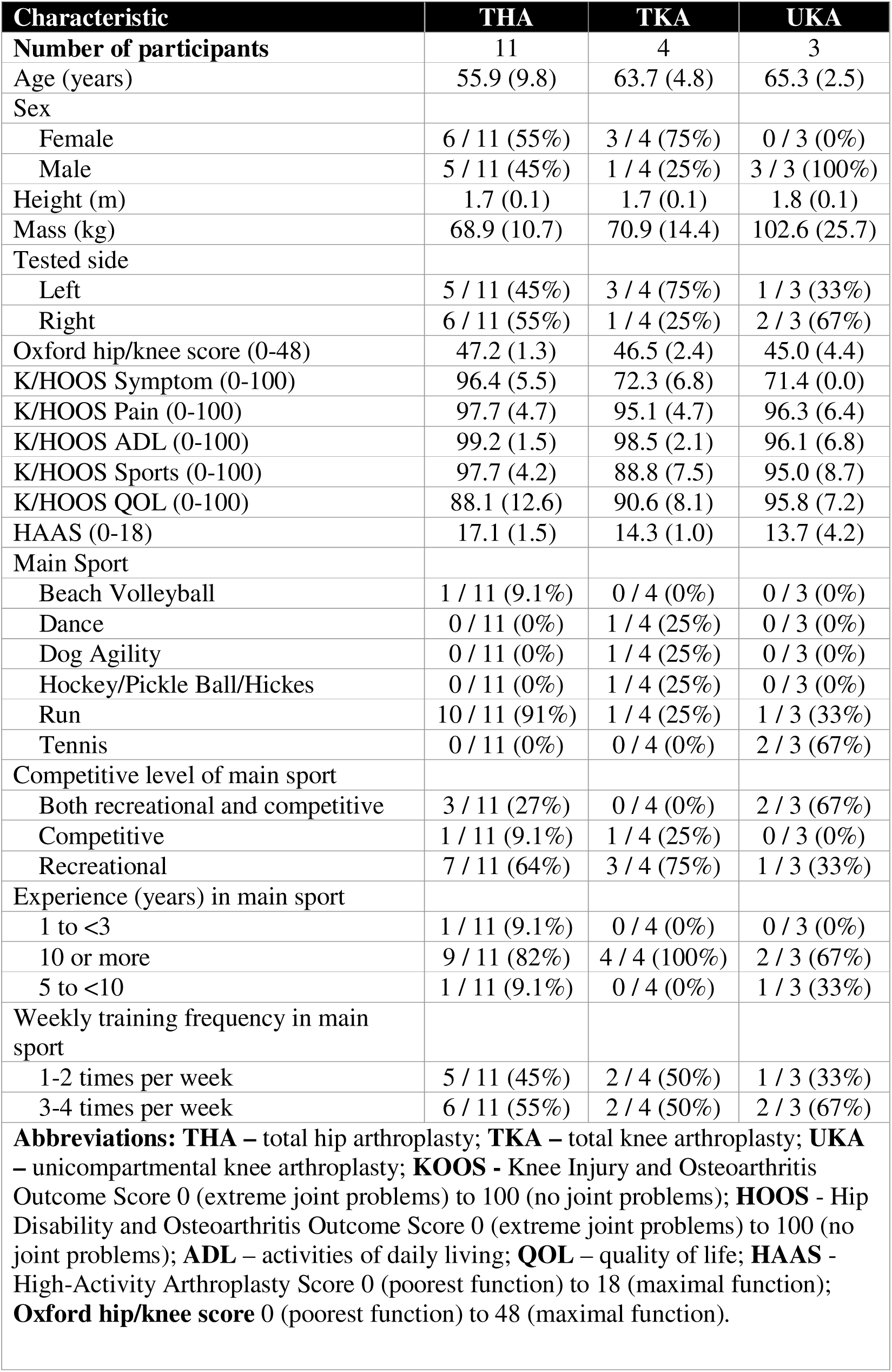
Demographic characteristics of participants with a joint replacement.

**Table 3.**
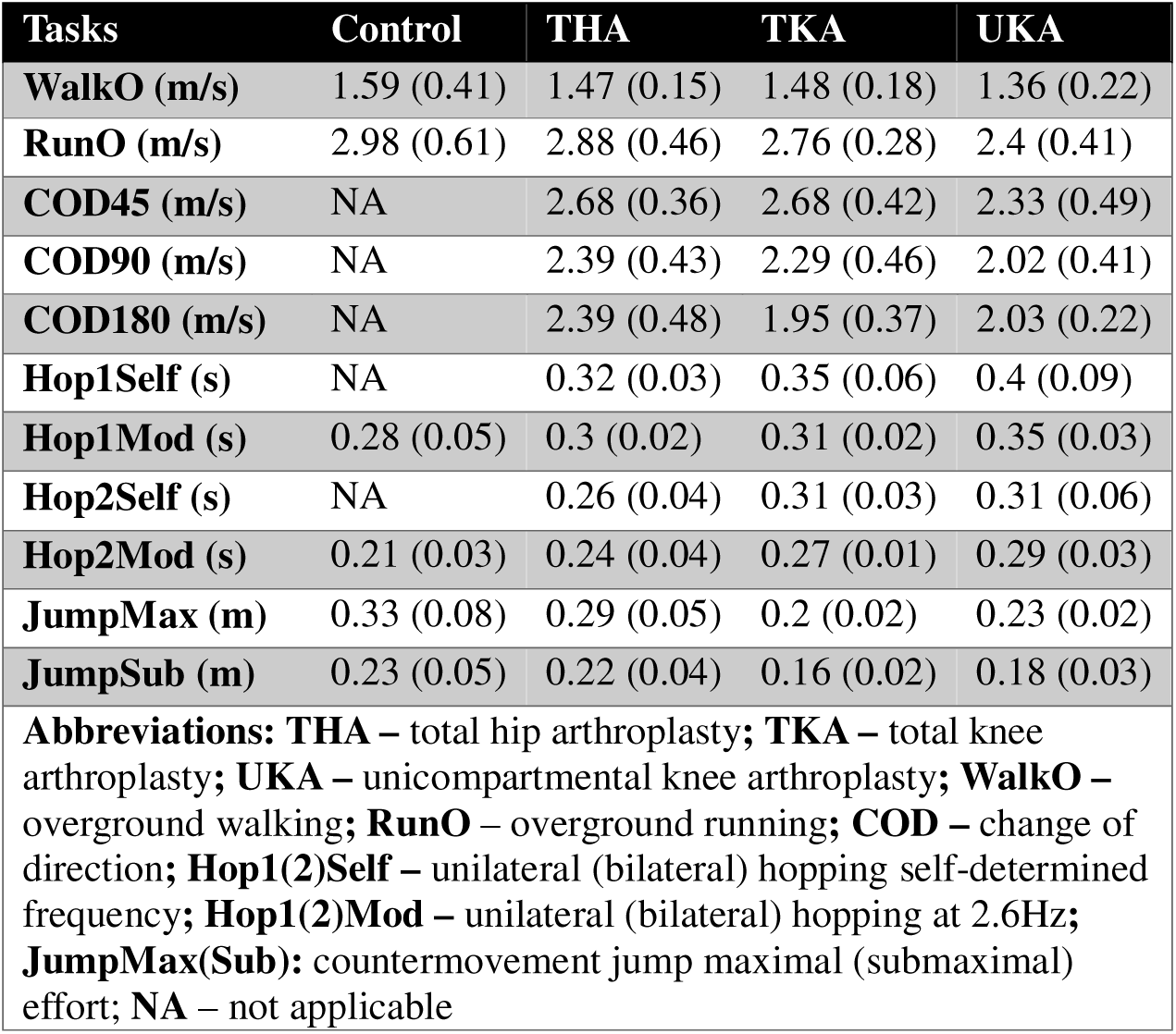
Mean (one standard deviation) of the intensity of the task performed.

| Tasks | Control | THA | TKA | UKA |
| --- | --- | --- | --- | --- |
| **WalkO (m/s)** | 1.59 (0.41) | 1.47 (0.15) | 1.48 (0.18) | 1.36 (0.22) |
| **RunO (m/s)** | 2.98 (0.61) | 2.88 (0.46) | 2.76 (0.28) | 2.4 (0.41) |
| **COD45 (m/s)** | NA | 2.68 (0.36) | 2.68 (0.42) | 2.33 (0.49) |
| **COD90 (m/s)** | NA | 2.39 (0.43) | 2.29 (0.46) | 2.02 (0.41) |
| **COD180 (m/s)** | NA | 2.39 (0.48) | 1.95 (0.37) | 2.03 (0.22) |
| **Hop1Self (s)** | NA | 0.32 (0.03) | 0.35 (0.06) | 0.4 (0.09) |
| **Hop1Mod (s)** | 0.28 (0.05) | 0.3 (0.02) | 0.31 (0.02) | 0.35 (0.03) |
| **Hop2Self (s)** | NA | 0.26 (0.04) | 0.31 (0.03) | 0.31 (0.06) |
| **Hop2Mod (s)** | 0.21 (0.03) | 0.24 (0.04) | 0.27 (0.01) | 0.29 (0.03) |
| **JumpMax (m)** | 0.33 (0.08) | 0.29 (0.05) | 0.2 (0.02) | 0.23 (0.02) |
| **JumpSub (m)** | 0.23 (0.05) | 0.22 (0.04) | 0.16 (0.02) | 0.18 (0.03) |
| **Abbreviations:** **THA** – total hip arthroplasty; **TKA** – total knee arthroplasty; **UKA** – unicompartmental knee arthroplasty; **WalkO** – overground walking; **RunO** – overground running; **COD** – change of direction; **Hop1(2)Self** – unilateral (bilateral) hopping self-determined frequency; **Hop1(2)Mod** – unilateral (bilateral) hopping at 2.6Hz; **JumpMax(Sub)**: countermovement jump maximal (submaximal) effort; **NA** – not applicable |  |  |  |  |

### Load ranking between tasks

For the dependent variable of HCF, there was a significant movement-by-group interaction effect (F [36, 860.09] = 2.24, P < 0.001), and a main effect of movement task (F [18, 860.09] = 73.28, P < 0.001). There was no main effect of group (F [2, 15.30] = 0.28, P = 0.761). For people with THA, HCF was statistically different from walking for 12 of 18 movement tasks (Figure 1a). Maximal and submaximal CMJs and all slope walking conditions resulted in HCFs like overground walking (Figure 1a). All COD tasks, running, and unilateral hopping resulted in >1.5 times the HCF magnitude found in walking (Figure 1a). COD45 resulted in the greatest HCF magnitude (8.38 [95%CI 7.70 to 9.06] BW), whilst bilateral hopping resulted in the least HCF magnitude (point estimates between 2.70 and 3.06 BW) (Figure 1a).

**Figure 1.**
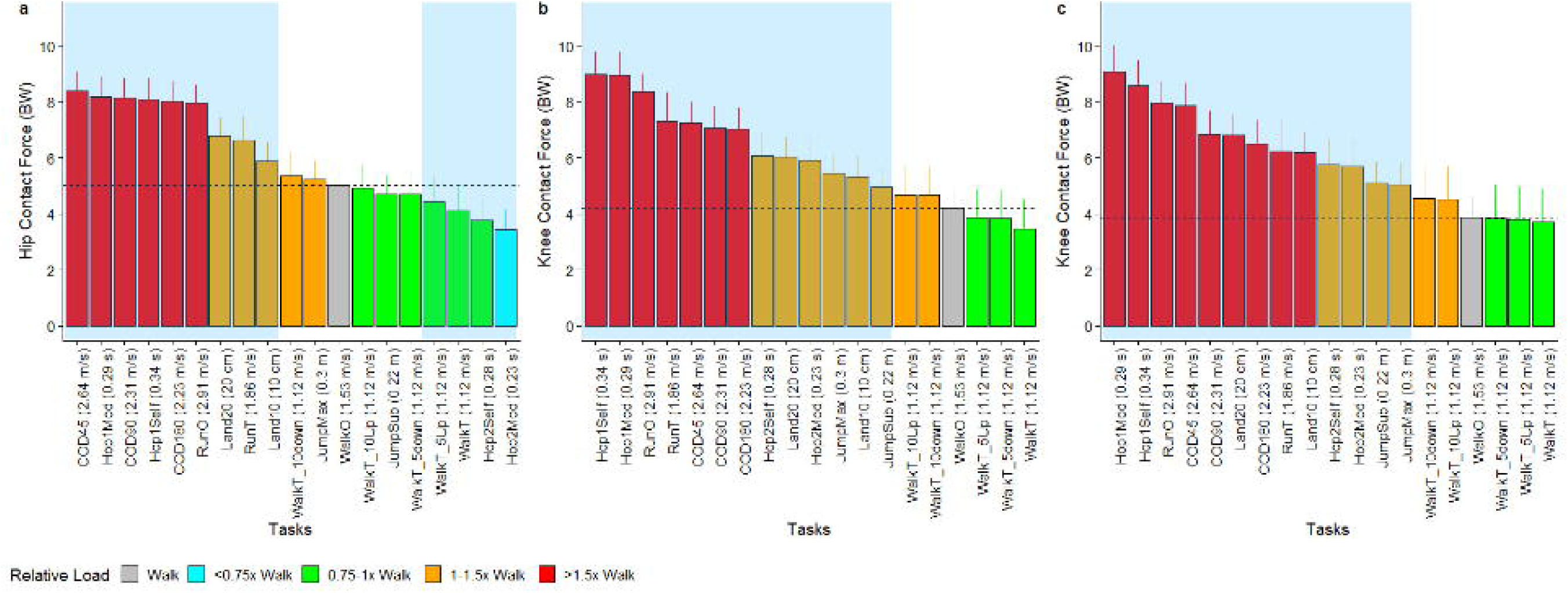
Predicted (95% confidence interval) of peak. (a) resultant hip contact force in people with a total hip arthroplasty, (b) resultant knee contact force in people with a total knee arthroplasty or (c) unicompartmental knee arthroplasty, across different activities. Dashe horizontal line represents the peak force reported in overground walking.

For the dependent variable of KCF, there was a significant movement-by-group interaction effect (F [36, 860.24] = 4.34, P < 0.001), and a main effect of both movement tasks (F [18, 860.25] = 133.66, P < 0.001) and group (F [2, 15.83] = 4.71, P = 0.025). For people with a TKA or a UKA, KCF was statistically greater in 13 out of 18 movement tasks compared to overground walking (Figure 1b, c). All five treadmill walking tasks resulted in peak KCF similar to overground walking in people with a TKA or UKA (Figure 1b, c). Across both TKA and UKA, unilateral hopping (point estimate between 8.56 to 9.07 BW) and overground running (8.00 to 8.36 BW) were the top three movement tasks with the greatest KCF (Figure 1b, c).

### Load differences between groups

For walking, there was a significant main effect of group on HCF (F [1, 47.49] = 12.27, P < 0.001). There was no significant main effect of group on HCF in running (F [1, 38.48] = 1.87, P = 0.180), unilateral (F [1, 44.51] = 0.35, P = 0.559) and bilateral hopping (F [1, 50.48] = 0.06, P = 0.810) at 2.6Hz, and CMJ maximal (F [1, 46.61] = 1.60, P = 0.212) (Figure 2a). Participants with a THA walked with 1.58 (95%CI 0.69 to 2.46) BW smaller HCF compared to controls (Figure 2a). There was no significant main effect of group on KCF for walking (F [2, 41.06] = 2.17, P = 0.127), for running (F [2, 38.41] = 0.23, P = 0.799), unilateral (F [2, 41.56] = 2.45, P = 0.099) and bilateral hopping at 2.6Hz (F [2, 51.93] = 1.16, P = 0.321), and CMJ maximal (F [2, 41.71] = 0.07, P = 0.934) (Figure 2b).

**Figure 2.**
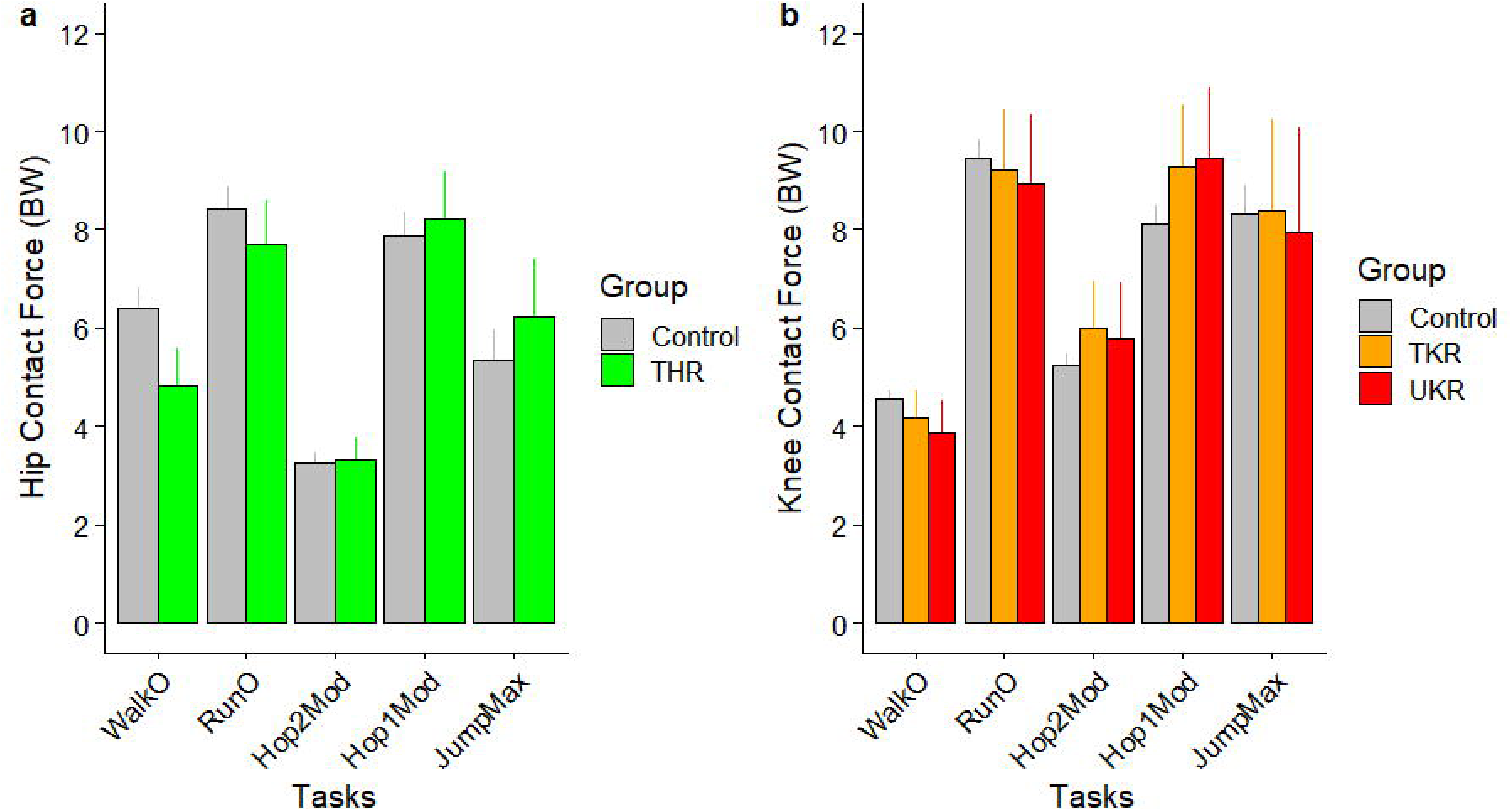
Predicted (95% confidence interval) of peak. (a) resultant hip contact force in people with a total hip arthroplasty vs healthy control, (b) resultant knee contact force in people with a total knee arthroplasty or unicompartmental knee arthroplasty vs healthy control. Abbreviations: WalkO – overground walking; RunO – overground running; COD – change of direction; Hop1(2) Mod – unilateral (bilateral) hopping at 2.6Hz; JumpMax: countermovement jump maximal effort.

## Discussion

Knowledge of the different load rankings across different PAs can help guide PA prescription, and the 3D force curves can be used for simulation implant testing [22]. In high-functioning participants after THA, COD45 produced the highest peak hip loading, whereas in participants after TKA/UKA, unilateral hopping and running produced the highest peak knee loading. These findings show that “high-impact” tasks vary substantially in peak joint loading rather than forming a single category.

Our modelled contact forces were slightly greater than in-vivo contact forces during walking, but greater during running [13, 14]. One reason could be that our overground walking and running speeds were around 0.4 m/s and around 1.0 m/s greater than Bergmann’s average speeds, respectively. Altai et al. reported greater HCF and KCF by 1.26BW and 0.84 BW, respectively, when increasing running speed by 1.2 m/s from a baseline of 3.0m/s [16]. The smaller in-vivo HCF and KCF during running may also be due to prior studies investigating non high-functioning patients who may not be participating in the activities that they are being assessed [13, 14]. This could have resulted in more cautious running patterns in participants from Bergmann et al [13, 14], like a reduced step length. Previous studies have reported that a 10% greater than preferred step length during running increases hip and knee energy absorption by 46 to 79% [23].

However, our modelled forces reflected values reported in prior in-silico modelling literature. Saxby et al. [24] reported peak KCFs of 2.83 BW, 7.83 BW, and 8.47 BW for walking, running, and 45° side-step cuts. Hip contact forces during slow to fast runs have been reported to be between 7.5 and 10.0 BW (speed range of 0.8–3.6 m/s) [25, 26]. Our HCF and KCF across running, hopping and CMJ were similar to those of healthy controls [16]. However, HCF during walking was lower than Altai et al [16], even after adjusting for speed and age. This could be due to the use of different scaling and musculoskeletal modelling algorithms [27]. Collectively, our results suggest that the present study cohort represented participants who performed high-impact PA more like healthy participants than patients after joint arthroplasty.

There is consistency in the present study, alongside the wider literature [16, 28], that unilateral hopping and running are activities that place the highest HCF. Previous studies have reported that femoral neck strain has a strong association with hip contact force magnitude (R^2^ = 0.80), where strain is approximately 745 times the hip contact force magnitude [29]. This suggests that unilateral hopping and running can place close to 6000 µε at the femoral neck [28]. However, such a load may be worrying, given that it is close to the elastic strain limit of bone [30]. Interestingly, the relationship between hip contact force and femoral neck strain varied from an R^2^ of 0.63 to 0.96, suggesting that this relationship may not be linear [31]. This could explain why a _prior study reported peak femoral neck strains in running < 3500 µ_ε[32].

The present results agree with clinical trials of the osteogenic potential of hopping-based exercises [9, 33]. On the one hand, unilateral hopping would provide sufficient osteogenic stimulus to increase bone density at the femoral neck in a person with a TKA. On the other hand, the same activity creates loads that far exceed the load magnitudes used for knee implant wear tests [34]. A previous study reported that a KCF of ∼10kN resulted in a tibial insert wear rate of 69 mm^3^/year, compared to a force of 2.2kN, which produced a wear rate of 14 mm^3^/year [35]. This increase in wear rate has been estimated to increase the risk of knee revision surgery from 0.1% to 0.5%, which the authors have also suggested may not be a clinically meaningful increase [35]. This is because the most significant cause of knee implant failure, aseptic loosening, has a risk of 1.1% [35].

A new finding from the present study is that COD also places joint loads on the hip comparable to those of unilateral hopping and running. This is highly relevant given that many social sports, such as racquet sports, require frequent COD [36]. Interestingly, COD tasks placed ∼2 BW less load on the knee implant but resulted in similar hip joint contact forces compared to unilateral hopping and running. An advantage of COD for people with a THA is that its multidirectional nature creates non-habitual bone strain patterns at the femoral neck that enhance the anabolic bone response [29].

The clinical acceptability of a heightened joint contact force will depend on the volume and variety of physical activity performed [37]. High joint load activities may not increase wear if they are undertaken for low periods of time [37]. For example, in post-menopausal women, 1 min of running a day is associated with better bone health [38]. Assuming a cadence of 180 steps/min, 65 700 high joint loading steps will be undertaken annually. In veteran football, running and COD may contribute up to 1160 steps within a 90 minute match[39, 40]. Based on the commonly used 1.1 million steps undertaken per year for wear simulation testing [41], high joint load steps may comprise ∼5% the annual steps undertaken, if a person ran daily for 1 min or played football once per week. The impact of different volumes and types of physical activities on implant longevity needs to be evaluated in future research.

The present study has several limitations. First, we did not have sufficient sample size to evaluate implant-level design–specific differences (e.g., brand/type) and their effects on joint loading. However, previous studies have found that joint kinetics are less influenced by implant type [42, 43]. Second, we did not incorporate different muscle recruitment strategies in our musculoskeletal models, which could further influence joint loading profiles. Lastly, given the small TKR and UKR sample sizes, subgroup estimates should be interpreted as exploratory, and confidence intervals may be relatively wide, indicating greater uncertainty around some estimates of impact activities.

## Conclusions

The present study provides a comparative analysis of joint loading data to the hip and knee in people with joint arthroplasty surgery during a variety of low– and high-impact PAs. Our joint load rankings could be used by researchers and clinicians for exercise prescription and PA counselling after a joint arthroplasty, particularly in the design of human clinical trials. Also, our data could be used for computational simulation of joint implants, which is an immediate plan for our research group.

## Ethics approval and consent to participate

All participants provided written informed consent, and the study received approval from the Health Research Authority Research Ethics Committee (IRAS 327418).

## Data Availability

All data produced in the present study are available upon reasonable request to the authors

## Acknowledgements

Not applicable

## Authors’ contributions

**Conceptualization,** BL, AS, LG, SMc, SM, NC; **Data Curation,** BL, JH, AS, ZA, WG; **Formal Analysis,** BL, JH, AS, ZA; **Funding Acquisition,** BL, AS, LG, SMc, WG, SM, NC. **Investigation,** BL, JH, AS, ZA, NC; **Methodology,** BL, AS, LG, SMc, WG, SM, NC; **Project Administration,** BL, ZA, NC.; **Resources,** BL, NC, ZA; **Software,** BL, JH, AS, ZA, WG; **Supervision,** BL, ZA, SM, NC; **Validation,** BL, JH, AS, ZA, WG, SMc; **Visualization,** BL, WG; **Writing –Original Draft,** BL, JH, AS, ZA, WG; **Writing – Review & Editing,** All Authors; **All authors** have read and approved the final version of the manuscript.

