## Supplementary Material for "Ranking hip and knee joint contact forces during high-impact activities in high-functioning adults after hip or knee arthroplasty"

**Journal category:** Original article

*Corresponding author

### Marker placement for Safejoints participants

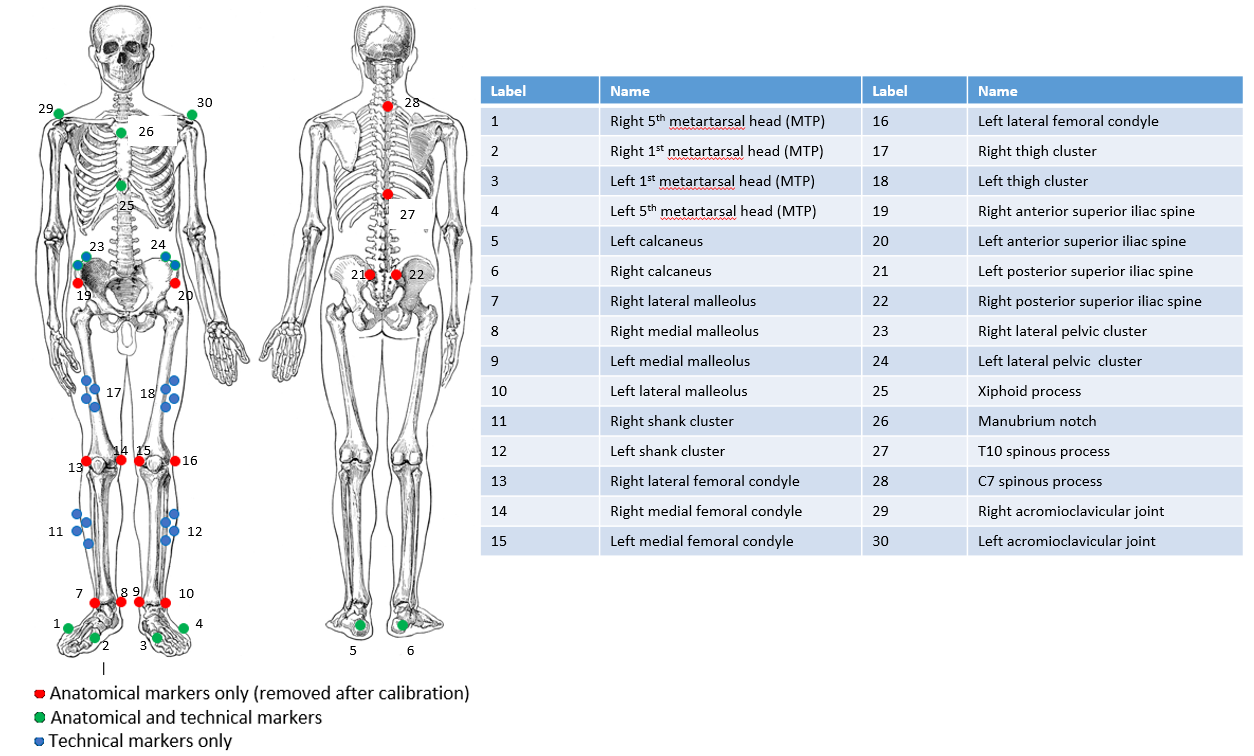

Figure SM 1. Marker template used for Safejoint participants

### Methodology summary

Table SM 1. Brief methodologies of included studies for healthy adults

|  | Altai et al. (Altai et al., 2024) |
| --- | --- |
| Country | England |
| Inclusion criteria | 40 healthy adults, free from injuries or pain |
| Camera system | 16 cameras (Vicon, UK) |
| Experimental protocol | All – 3 successful trials each, overground, shod (own pair of exercise shoes)  Wak – self-determined pace  Run – self-determined pace  CMJ – maximal and medium (50%) effort, arms abducted “T” pose, self-selected depth  Hopping – vertical bilateral and unilateral (dominant, side used to kick ball), at 2.6Hz for 10s |
| Camera sampling frequency | 200 Hz |
| Force platform system | 2 force plates (Kistler, Switzerland) |
| Force sampling frequency | 2000Hz |
| Filtering | Markers – 18 Hz (2^nd^ Order, Butterworth, Zero lag)  GRF – 50 Hz (2^nd^ Order, Butterworth, Zero lag) |

### Coordinate systems to resolve hip and knee forces

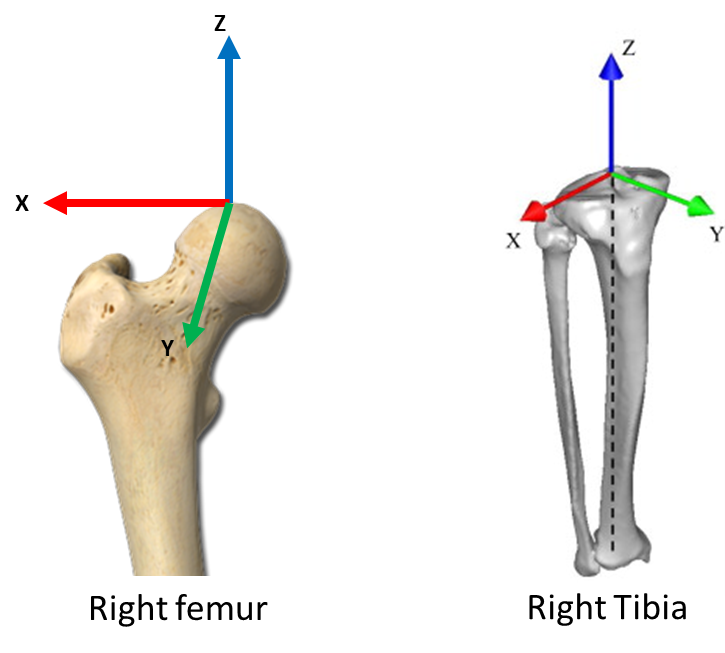

### Plots of resultant joint contact force waveforms

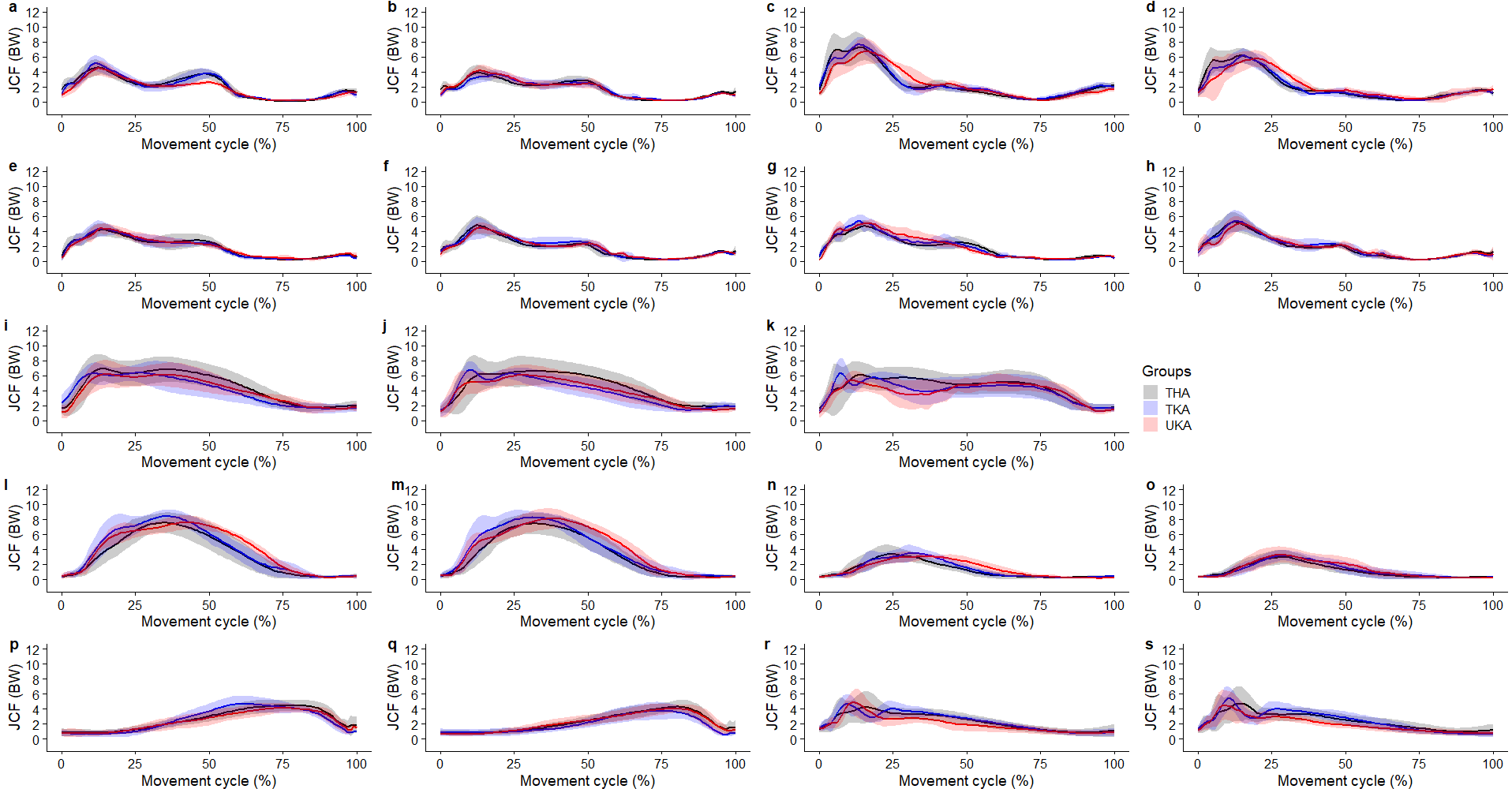

Figure SM 2. Mean with error clouds as one standard deviation of the resultant **hip contact force** over a movement cycle. Subplots: **a)** overground **(b)** treadmill self-paced walking; **(c)** overground and **(d)** treadmill self-paced running; treadmill walking **(e)** 5° upslope, **(f)** 5° downslope, **(g)** 10° upslope, **(h)** 10° downslope; (i) change of direction 45°, **(j)** 90°, **(k)** 180°; **(l)** unilateral hopping self-paced and **(m)** 2.6Hz frequency, **(n)** bilateral hopping self-paced and **(o)** 2.6Hz frequency; **(p)** maximal and **(q)** submaximal, **(r)** bilateral landing from 10 cm step and **(s)** 20 cm step. **Abbreviations:** THA – total hip arthroplasty, TKA- total knee arthroplasty, UKA – unicompartmental knee arthroplasty.

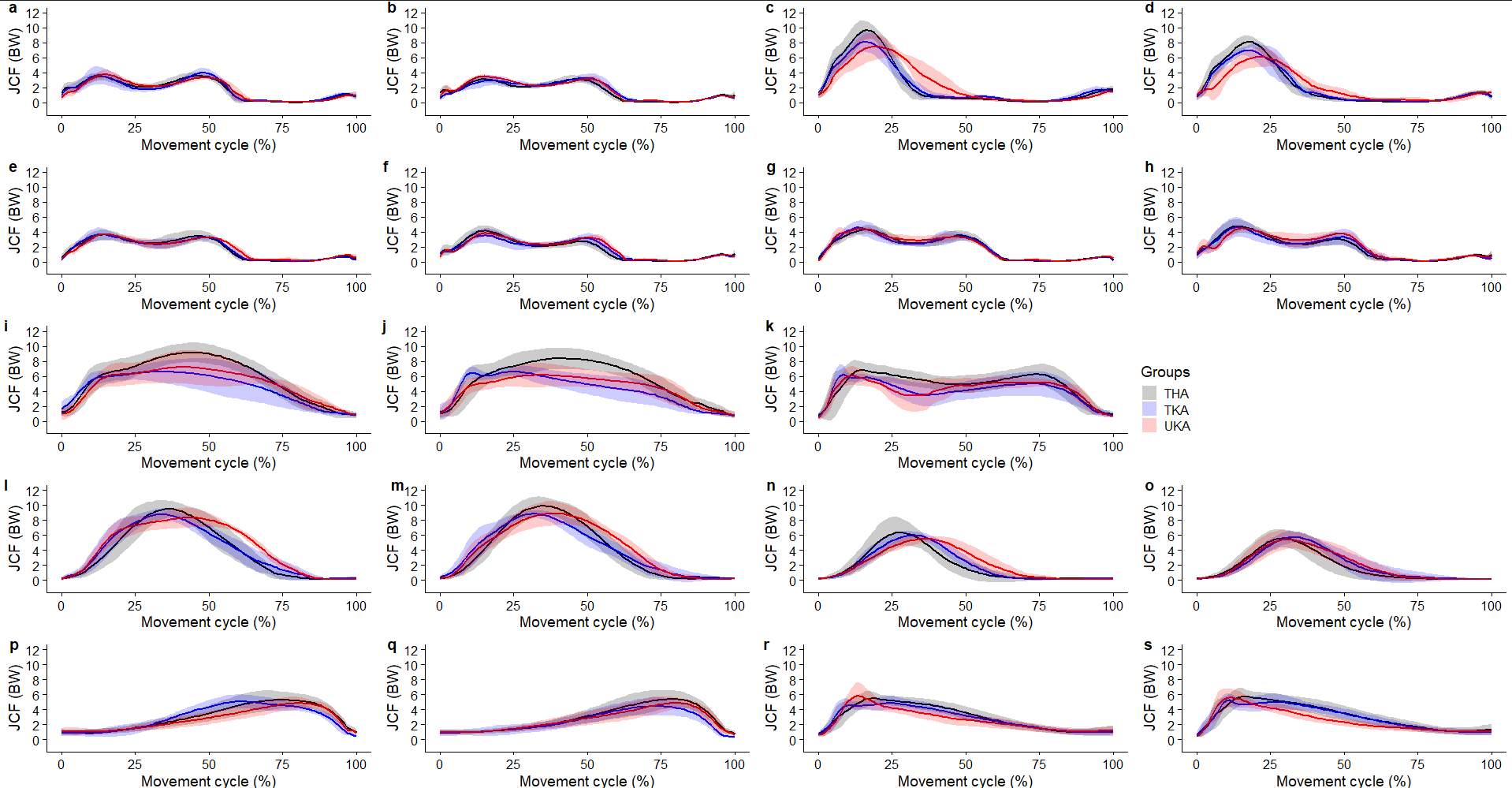

Figure SM 3 Mean with error clouds as one standard deviation of the resultant **knee contact force** over a movement cycle. Subplots: **a)** overground self-paced walking and **(b)** running; **(c)** treadmill self-paced walking and **(d)** running; treadmill walking **(e)** 5° upslope, **(f)** 5° downslope, **(g)** 10° upslope, **(h)** 10° downslope; (i) change of direction 45°, **(j)** 90°, **(k)** 180°; **(l)** unilateral hopping self-paced and **(m)** 2.6Hz frequency, **(n)** bilateral hopping self-paced and **(o)** 2.6Hz frequency; **(p)** maximal and **(q)** submaximal, **(r)** bilateral landing from 10 cm step and **(s)** 20 cm step. **Abbreviations:** THA – total hip arthroplasty, TKA- total knee arthroplasty, UKA – unicompartmental knee arthroplasty.

### Plots of individual axis components of joint contact force waveforms

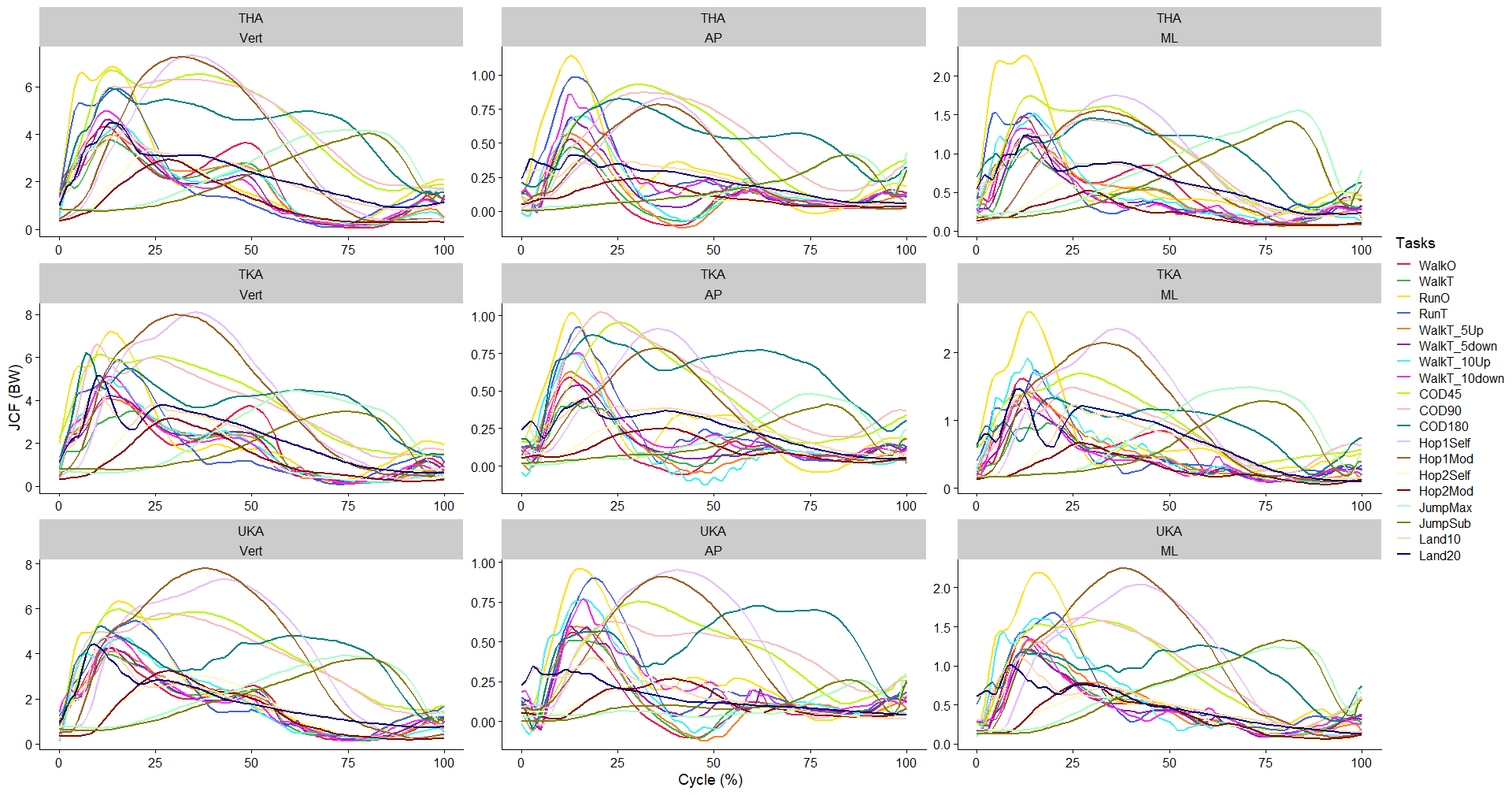

Figure SM 4 Mean resultant **hip contact force** over a movement cycle. **Abbreviations:** THA – total hip arthroplasty, TKA- total knee arthroplasty, UKA – unicompartmental knee arthroplasty, Vert – vertical force (+ compression), AP – anterior posterior force (+ posterior shear), ML – medial-lateral (+lateral force).

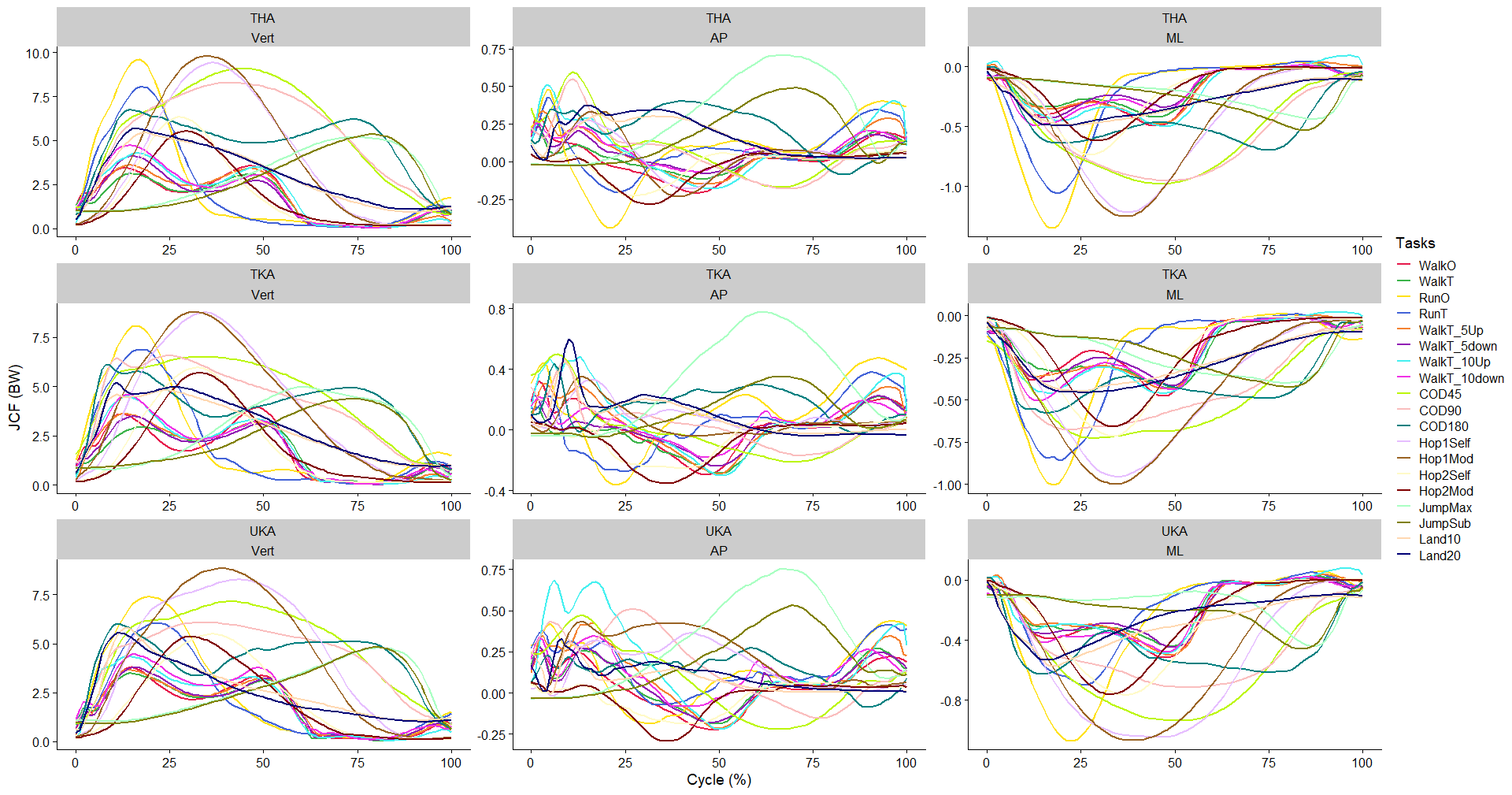

Figure SM 5. Mean resultant **knee contact force** over a movement cycle. **Abbreviations:** THA – total hip arthroplasty, TKA- total knee arthroplasty, UKA – unicompartmental knee arthroplasty, Vert – vertical force (+ compression), AP – anterior posterior force (+ posterior shear), ML – medial-lateral (+lateral force).

### Comparing modelled forces with Bergmann

**Link to download hip forces:** <https://orthoload.com/test-loads/standardized-loads-acting-at-hip-implants/>

**Link to download knee forces:** <https://orthoload.com/test-loads/standard-loads-knee-joint/>

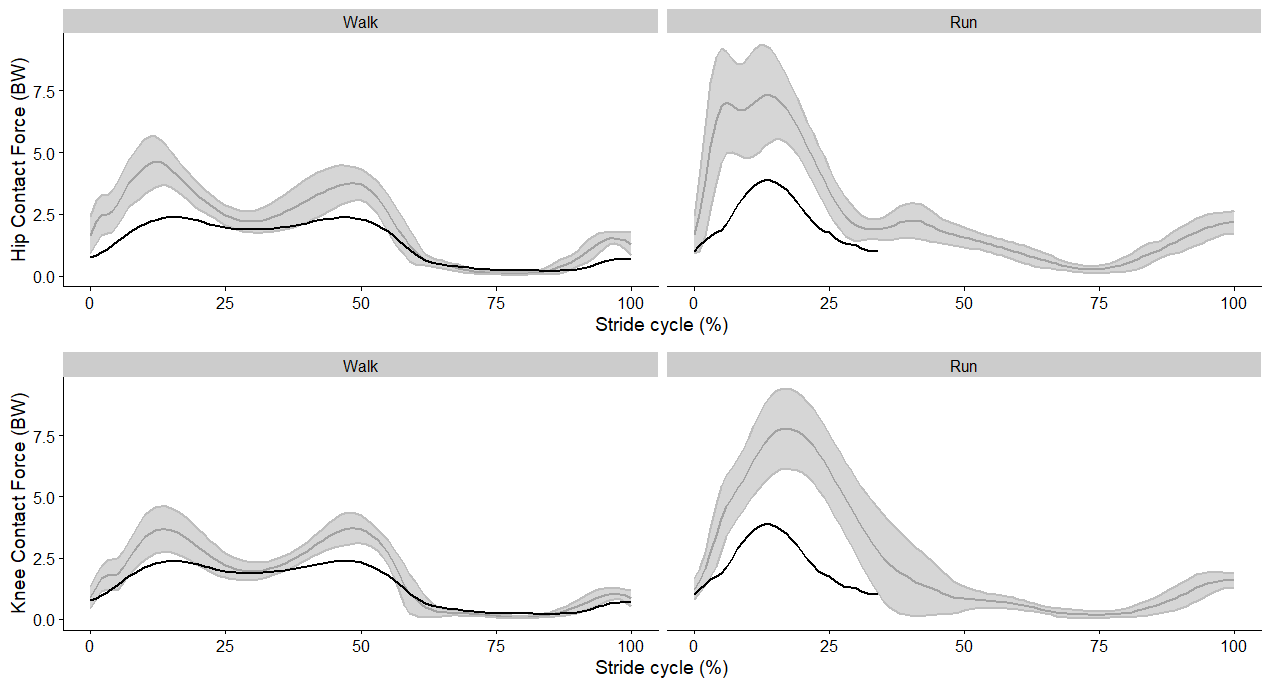

Figure SM 6. Normalised

### Comparing modelled muscle forces with electromyography for a single subject

Electromyography (EMG) sensors (Noraxon USA, 2 kHz) were attached unilaterally to the operated side to the following muscles: gluteus maximus, gluteus medius, vastus lateralis, biceps femoris, and soleus following SENIAM guidance (Hermens et al., 2000). Surface EMG signals were high-pass filtered at 50 Hz using a zero-phase lag, fourth-order Butterworth filter, rectified, low-pass filtered at 12 Hz, and amplitude normalised to the peak value within the movement cycle for each task (Table 1 in main manuscript). Normalising EMG to its peak value within a task scales its value to a range of 0 (no activation) and 1 (maximal activation). The force (N) for the same muscles was also scaled to a range of 0 (no force) and 1 (maximal force).

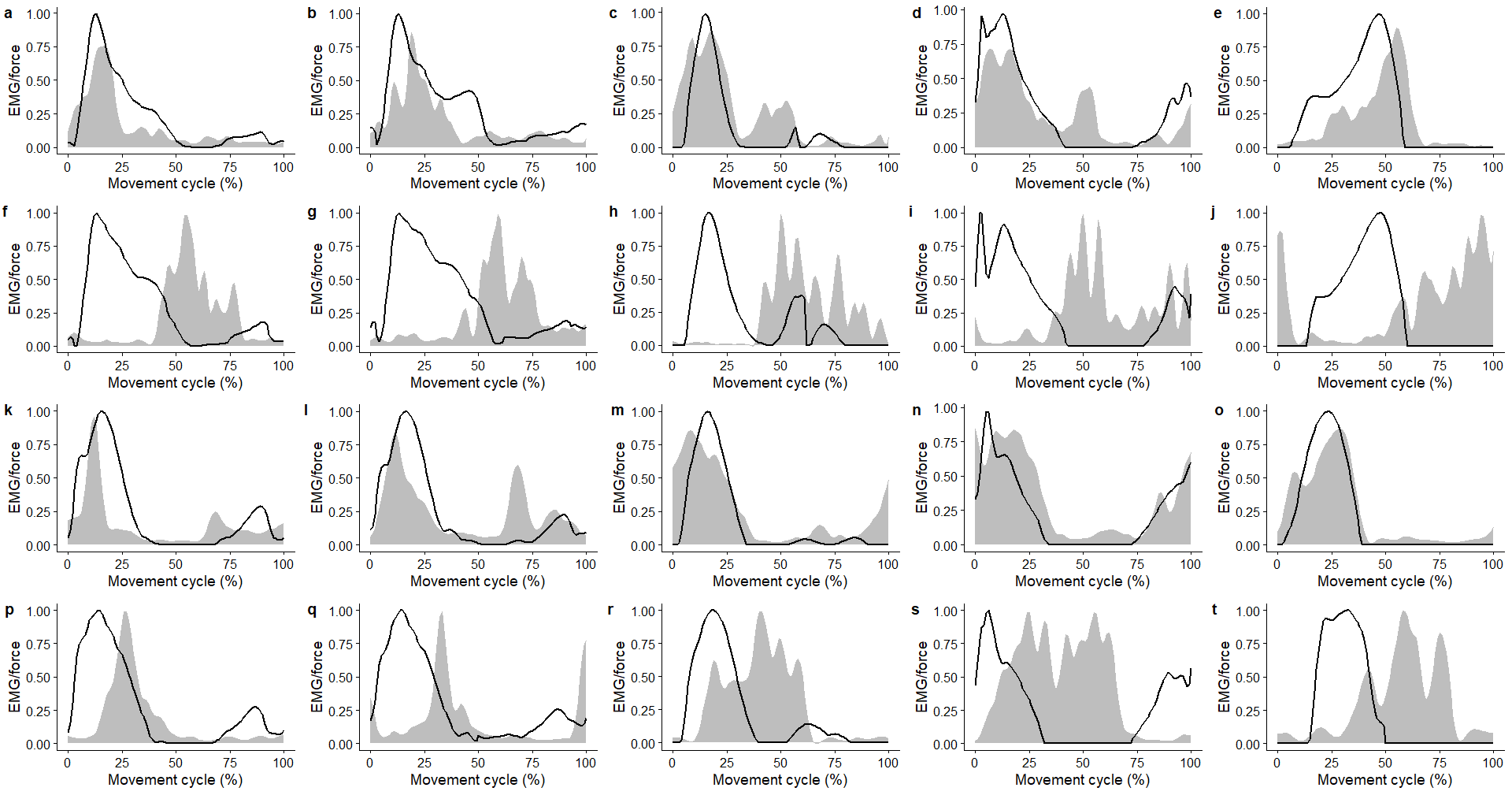

Figure SM 7. Plots of relative electromyography activation (grey area) against modelled muscle force (black line) for **(1^st^ row)** overground walking, **(2^nd^ row)** treadmill walking, **(3^rd^ row)** overground running, **(4^th^ row)** treadmill running, of the following muscle (left to right): **(1^st^ column)** Gluteus Maximus, **(2^nd^ column)** Gluteus Medius, **(3^rd^ column)** Vastus Lateralis, **(4^th^ column)** Biceps Femoris, **(5^th^ column)** Soleus.

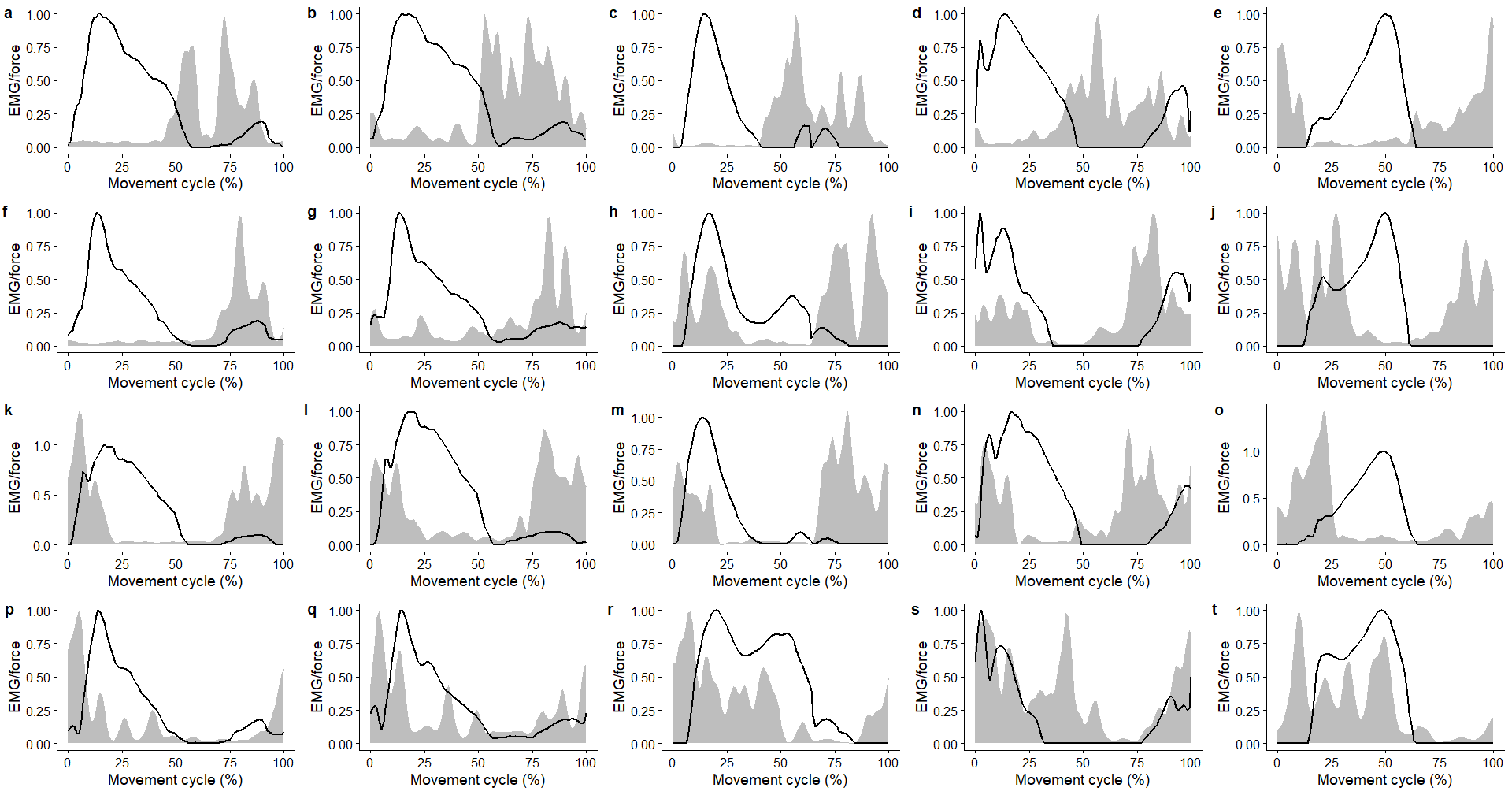

Figure SM 8. Plots of relative electromyography activation (grey area) against modelled muscle force (black line) for treadmill walking **(1^st^ row)** 5° upslope, **(2^nd^ row)** 5° downslope, **(3^rd^ row)** 10° upslope, **(4^th^ row)** 10° downslope, of the following muscle (left to right): **(1^st^ column)** Gluteus Maximus, **(2^nd^ column)** Gluteus Medius, **(3^rd^ column)** Vastus Lateralis, **(4^th^ column)** Biceps Femoris, **(5^th^ column)** Soleus.

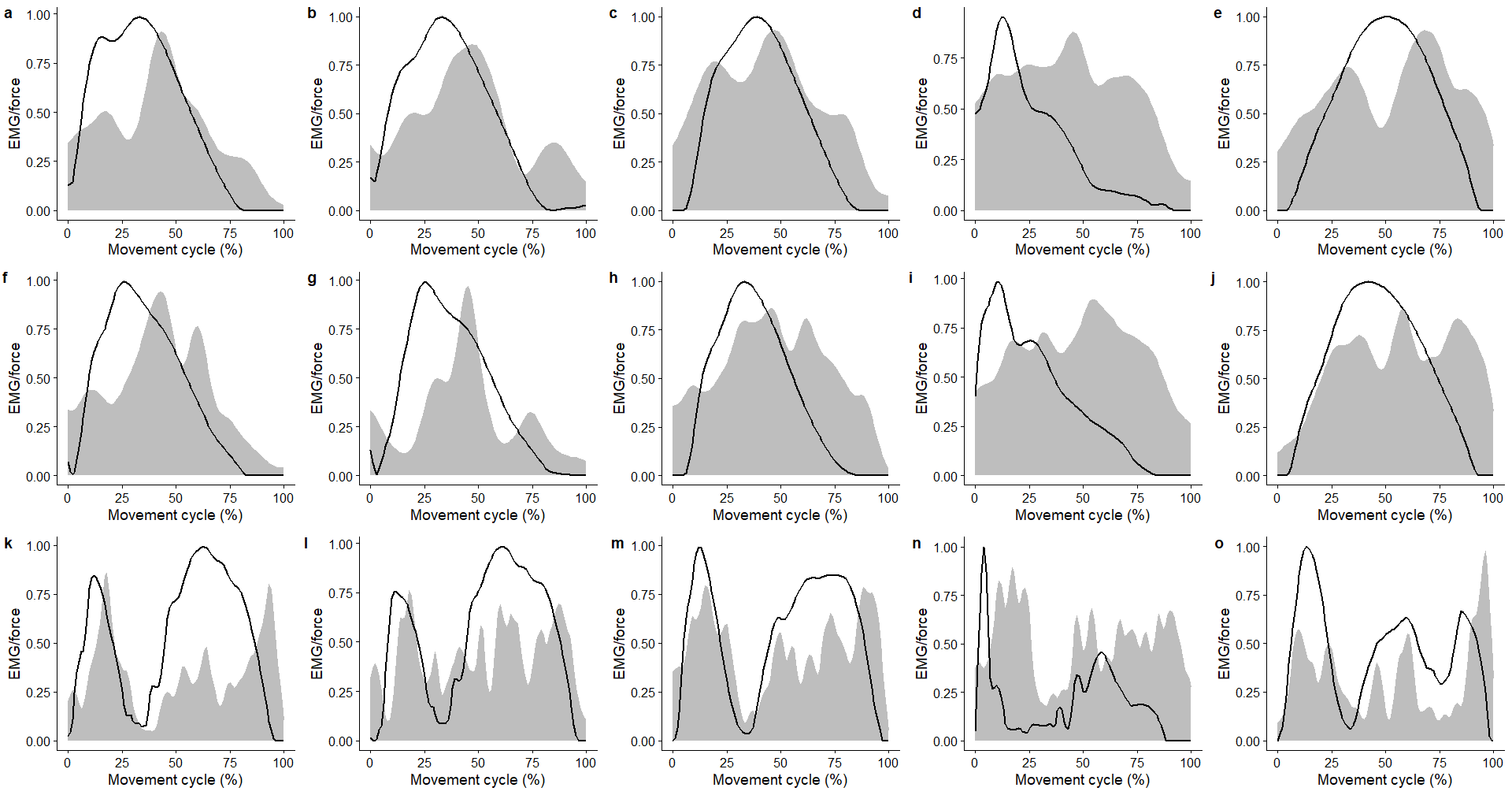

Figure SM 9. Plots of relative electromyography activation (grey area) against modelled muscle force (black line) for change of direction **(1^st^ row)** 45°, **(2^nd^ row)** 90°, **(3^rd^ row)** 180° of the following six muscles: **(1^st^ column)** Gluteus Maximus, **(2^nd^ column)** Gluteus Medius, **(3^rd^ column)** Vastus Lateralis, **(4^th^ column)** Biceps Femoris, **(5^th^ column)** Soleus.

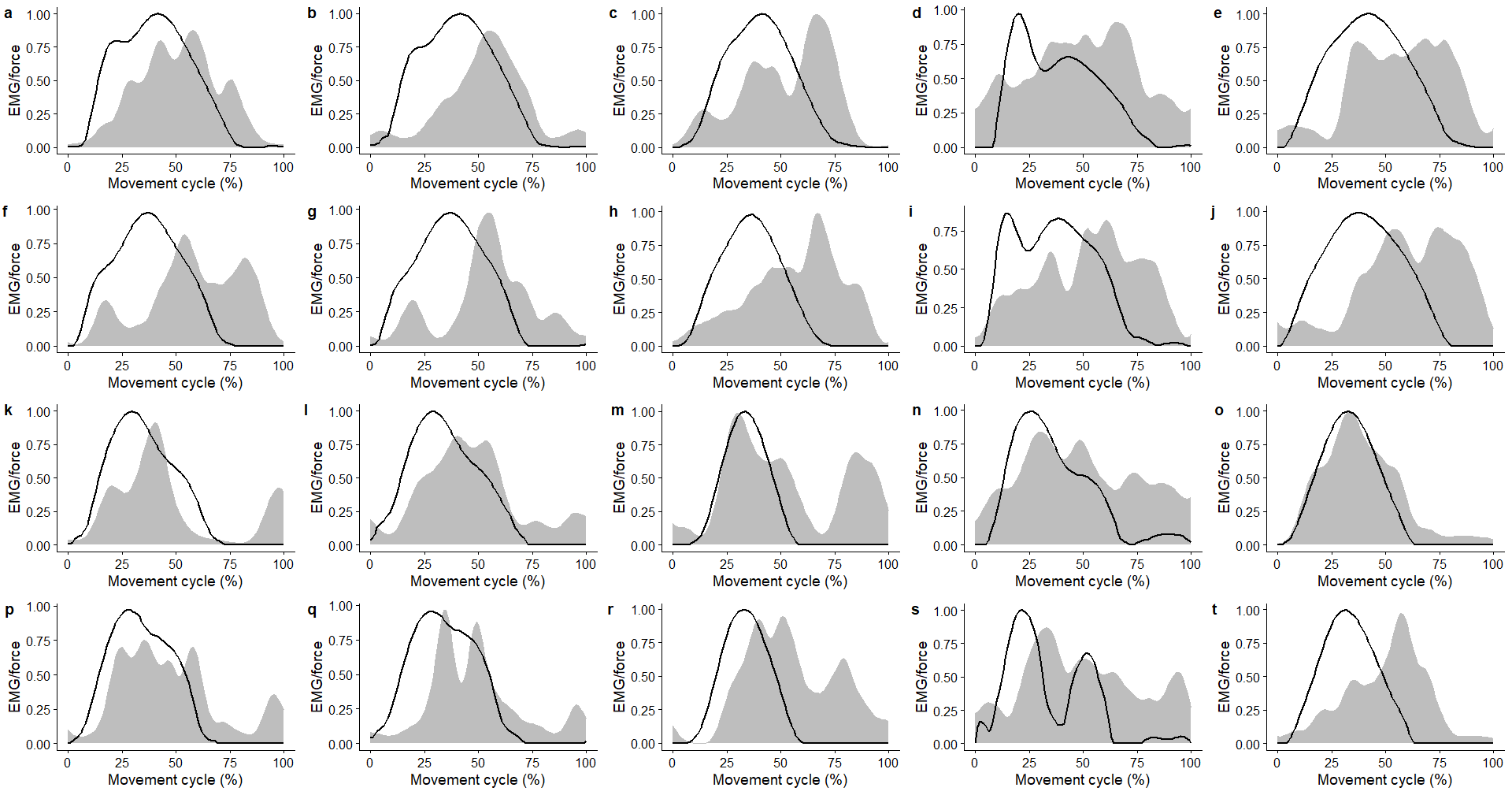

Figure SM 10. Plots of relative electromyography activation (grey area) against modelled muscle force (black line) for **(1^st^ row)** Unilateral hopping self-paced, **(2^nd^ row)** Unilateral hopping 2.6Hz, **(3^rd^ row)** bilateral hopping self-paced, **(4^th^ row)** bilateral hopping 2.6hz, of the following six muscles: **(1^st^ column)** Gluteus Maximus, **(2^nd^ column)** Gluteus Medius, **(3^rd^ column)** Vastus Lateralis, **(4^th^ column)** Biceps Femoris, **(5^th^ column)** Soleus.

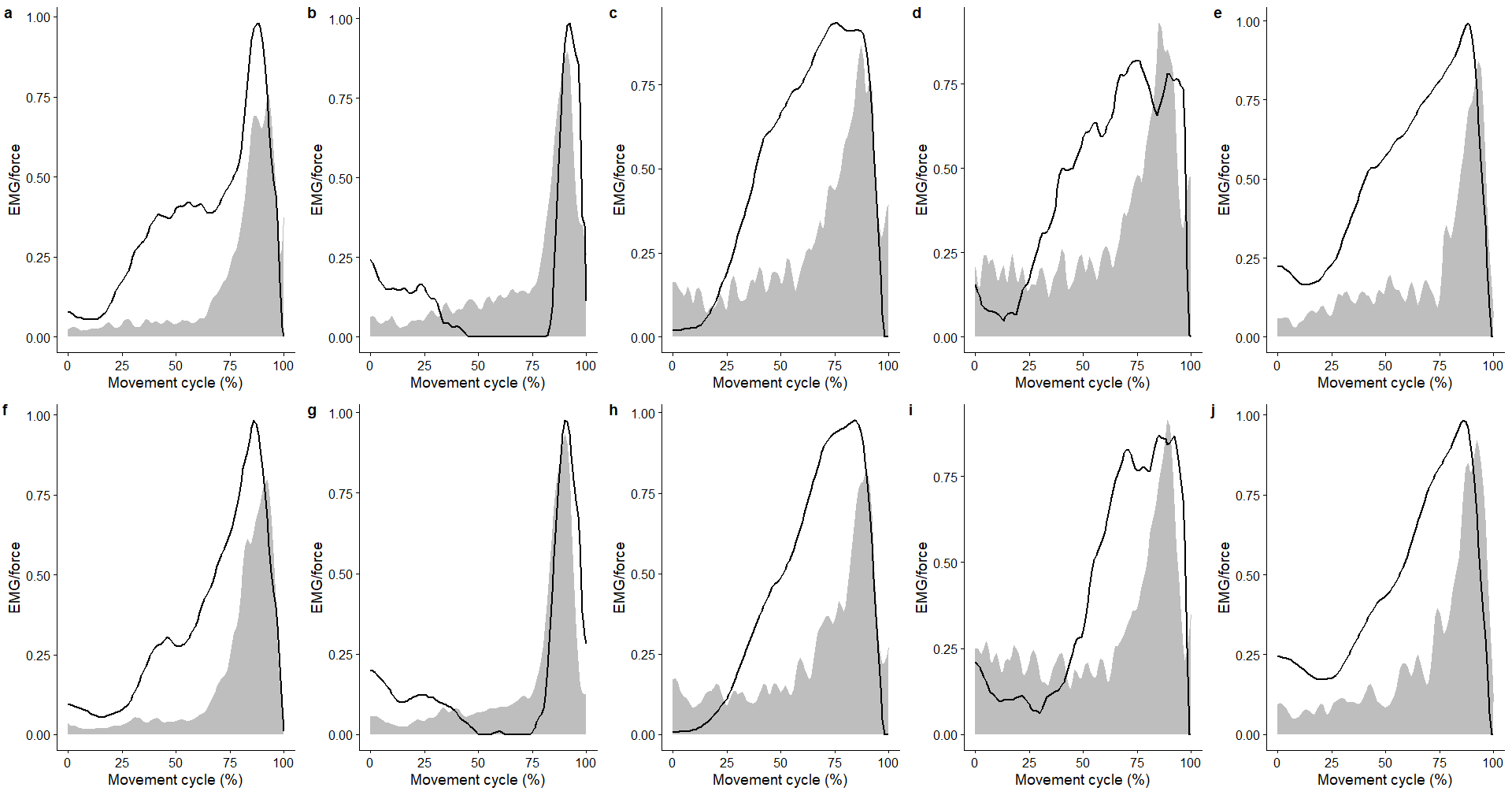

Figure SM 11. Plots of relative electromyography activation (grey area) against modelled muscle force (black line) for **(1^st^row)** maximal countermovement jump and **(2^nd^ row)** submaximal countermovement jump of the following six muscles: **(1^st^ column)** Gluteus Maximus, **(2^nd^ column)** Gluteus Medius, **(3^rd^ column)** Vastus Lateralis, **(4^th^ column)** Biceps Femoris, **(5^th^ column)** Soleus.

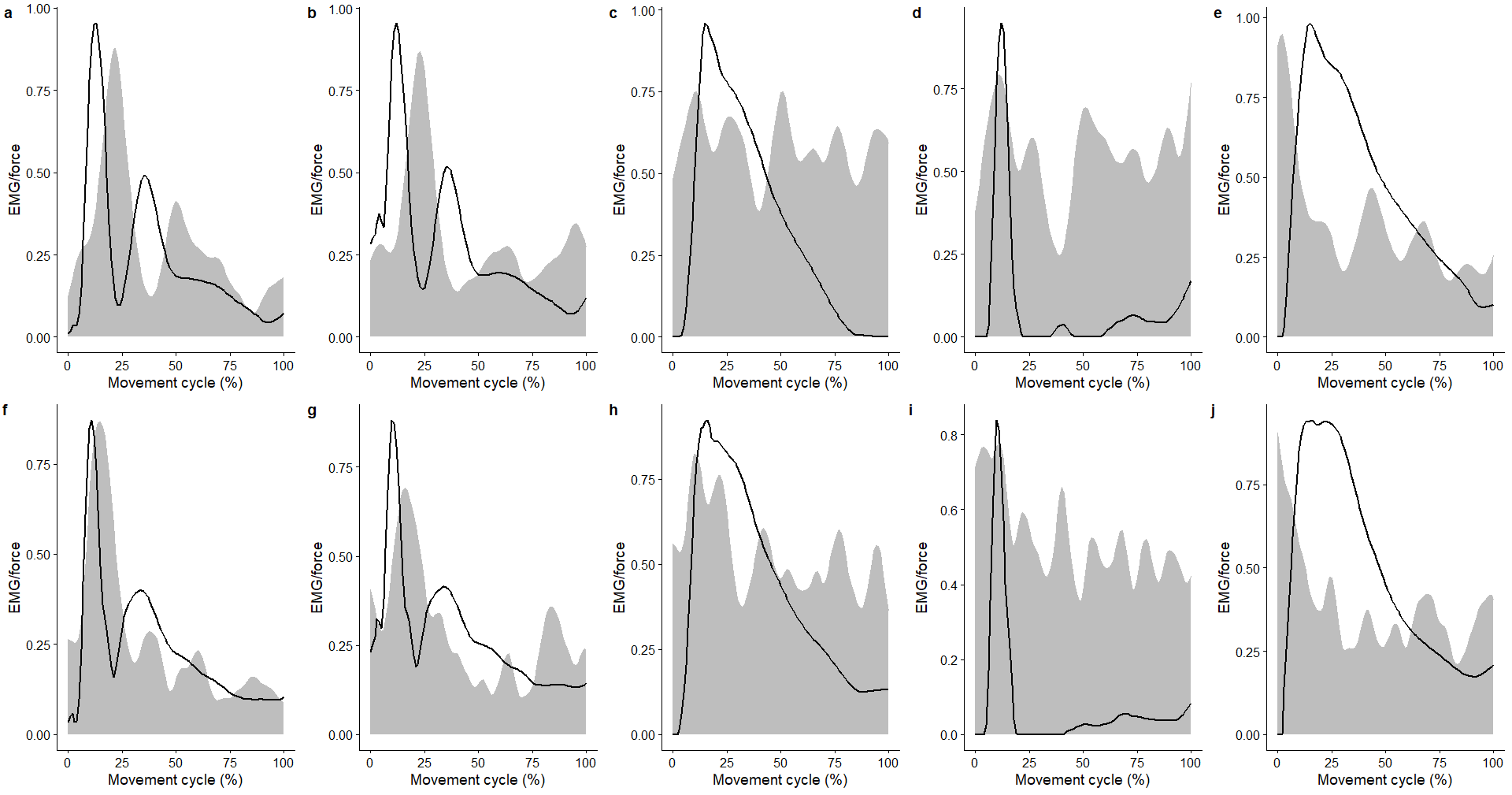

Figure SM 12. Plots of relative electromyography activation (grey area) against modelled muscle force (black line) for **(1^st^ row)** landing from a 10cm step and **(2^nd^ row) landing** from a 20cm step, of the following six muscles: **(1^st^ column)** Gluteus Maximus, **(2^nd^ column)** Gluteus Medius, **(3^rd^ column)** Vastus Lateralis, **(4^th^ column)** Biceps Femoris, **(5^th^ column)** Soleus.
